# Dynamics of psychological responses to Covid-19 in India: A longitudinal study

**DOI:** 10.1101/2020.06.12.20129791

**Authors:** Anvita Gopal, Anupam Joya Sharma, Malavika Ambale Subramanyam

**Affiliations:** Center for Cognitive and Brain Sciences, Indian Institute of Technology Gandhinagar, Gujarat 382 355, India; Social Epidemiology, Indian Institute of Technology Gandhinagar, Gujarat 382 355, India

**Keywords:** Longitudinal, India, stress, gender disparity, anxiety, depression, pandemic, mental health

## Abstract

To curb the spread of the novel coronavirus, India announced a nationwide lockdown on 24th March 2020 for 21 days, later extended for a longer time. This long period of lockdown greatly disrupted routine life and likely affecting citizens’ psychological well-being. The psychological toll of the pandemic on Indians is documented. However, no study has assessed whether the psychological toll changed over time due to repeated extensions of the lockdown. We followed up 159 Indian adults during the first two months of the lockdown to assess any change in their anxiety, stress, and depressive symptoms. Multilevel linear regression models of repeated observations nested within individuals, adjusted for socio-demographic covariates, showed that anxiety (**β**=0.81, CI: 0.03, 1.60), stress (**β**=0.51, CI: 0.32, 0.70), and depressive symptoms (**β**=0.37, CI: 0.13, 0.60) increased over time during the lockdown. This increase was higher among women than men independent of covariates. Individual resilience was negatively associated with the psychological outcomes. This suggests that the state needs to address the current mental health impacts of a long-drawn out lockdown and its long-term sequelae. Disproportionate burden on women needs immediate attention. Sustainable change requires addressing the root causes driving the gender inequalities in psychological distress during such crises.

## 1. Introduction

The novel coronavirus disease (COVID-19), which originated in China, was declared a public health emergency by WHO on January 30th, 2020 [^1^]. With a steep global increase in the number of infected persons, different countries took stringent measures to curb its spread, including nationwide “lockdowns.” The government of India called for a nationwide lockdown from the 25th of March, 2020 [^2^]. Citizens were mandated to stay at home and all major offices, malls, factories and schools were ordered to be shut down for 21 days [^2^]. However, it was extended until 3 May, with conditional relaxations [^3^]. While the lockdown was intended to curb the spread of the virus, it likely had a psychological impact on the citizens[^4–7^]. The restrictions on physical mobility due to the lockdown and measures of self-isolation caused major disruptions to routine lives as well as it hindered meeting of regular responsibilities--potentially affecting the physical and mental health of individuals. Recent studies have studied the higher levels of stress [^8,9^], anxiety [^8,10,11^], depression [^10,12,13^], and poorer quality of life [^9,10^] during the Covid-19 crisis in different populations. However, the extensions in the lockdown period in India led to longer restrictions on physical mobility and prolonged self-isolation measures, which could have increased the intensity of negative psychological outcomes among Indian adults, leading to a poorer quality of life, not just during the lockdown, but also after the crisis. Previous studies have shown that prolonged periods of isolation and limited mobility significantly impacted mental wellbeing [^14,15^] during crises. Further, such prolonged exposure to negative mental health outcomes could have adverse effects on physical health outcomes, such as sleep disorders[^16^] and health-related quality of life[^17^].

While mental health effects due to the lockdown are likely to be seen among a majority of Indian adults, the effect of a lockdown *extended over longer periods of time* might differ across vulnerable groups. For instance, the stress experienced by persons with limited adaptive capacity, fewer financial resources, lower social support, and pre-existing mental health conditions, might be higher than among those who do not share these characteristics. As the lockdown period increases, financially weaker individuals might face greater challenges in meeting the basic needs of their family. Moreover, the continued restrictions on physical mobility could place a greater burden on the social networks of vulnerable individuals, thus reducing access to social support over time and impacting their adaptability. Further, in a patriarchal society such as India, with a high prevalence of domestic abuse [^18^], the lockdown[^19^](especially limited mobility) potentially increases the risk of experiencing domestic abuse, prolonged exposure to which can worsen the mental health of women during this crisis.

Despite these risks, several resources that help with coping could be available to individuals. Previous studies have highlighted the role of social support in reducing anxiety and stress[^20,21^]. Recent studies focused on COVID-19 also support this [^11,22^]. In addition to social support, there could also be several individual-level resources such as resilience that could help individuals face adversity [^23^]. Resilience was found to help strengthen mental health and reduce the possibility of developing psychiatric morbidities, especially during the COVID-19 pandemic[^24,25^]. There is scant research on the mental health effects of such protective factors during the extended lockdown in India.

Reliably measuring the impact of lockdowns that extend over a long period of time requires a longitudinal study design. While several cross-sectional studies[^5–7^] have focused on psychological wellbeing during Covid-19 in India, we could not locate any study investigating the *change* in such psychological outcomes throughout the lockdown period. A longitudinal investigation helps establish temporal sequence and document trends while investigating how the adverse psychological outcomes change (if at all) over time during the lockdown. To address this gap in the literature, we conducted a longitudinal study in three phases to investigate the changes in three psychological outcomes viz, anxiety, stress, and depressive symptoms, during the lockdown in India. The research questions we addressed in our study were:

1. Do the levels of anxiety, stress, and depressive symptoms change during the lockdown among Indian adults, independent of their age, gender, income, educational qualification, place of residence, and history of mental health? We hypothesized that levels of anxiety, stress, and depressive symptoms will increase over time, independent of the covariates.
2. Do these changes, if any, in the levels of anxiety, stress, and depressive symptoms, differ by gender? Based on the large body of research highlighting gender disparities in the risk of anxiety, stress, and depression[^26^], we hypothesized that in a patriarchal society such as India, compared to men, women will have a greater increase in levels of anxiety, stress, and depression during the lockdown.
3. Are social supports available to an individual and the level of their personal resilience related to any changes in levels of their anxiety, stress, and depression? We hypothesized that higher greater social support and higher resilience will be related to lower levels of anxiety, stress, and depression, independent of all covariates.

## 2. Materials and Methods

### Recruitment of participants

We collected quantitative repeated measures data on psychological wellbeing during the lockdown via a set of four web-based surveys, which were administered in an intermittent manner to the same participants during 2 months of the COVID-19 lockdown in India. Online Google and Microsoft forms were circulated through social media platforms such as Facebook and LinkedIn to recruit a diverse pool of participants. This method of recruitment was suitable due to the restrictions on in-person interactions with strangers during the time of the lockdown in India, and efficient, given the ability to recruit a diverse sample of participants very quickly. In addition to posting the Google forms on social media, they were circulated among the social networks of the authors. Moreover, we requested the participants to further share the form among their peers to increase the size as well as the diversity of the sample. All the forms included a brief introduction describing the major objectives of the study. Additionally, all the participants were informed that their participation was completely voluntary.

We deployed our first survey on March 29th, during the first week of the lockdown. The online survey was open for 2 weeks. We received responses from 793 participants in this round (T1). However, only 561 of them shared their interest in participating in the subsequent surveys. We rolled out the second follow-up survey on April 14th, 2020, the third on 2nd May, and the final follow-up survey on the 24th of May. Since we recruited our participants through social media, the second (T2), third (T3), and the fourth (T4) surveys received responses from newer participants as well. Our analytical sample for the current study included only the 159 participants from India who voluntarily participated in all four follow-ups. However, we measured the outcomes of interest of the current study only at two time points. Data on anxiety and stress were collected during T1 and T4, while depressive symptoms were measured during T2 and T4.

### Response variables

#### Anxiety

We used the Generalized Anxiety Disorder-7 (GAD-7) scale to assess anxiety. The scale has been widely used with a demonstrated high reliability and validity[^27^]. The scale included seven items such as “Feeling nervous, anxious or on edge; Not being able to stop or control worrying” The responses were recorded on a 4-point Likert scale ranging from *never* (0) to *nearly every day* (3). The total score of GAD-7 ranged from 0 to 21. Greater score predicted higher levels of anxiety[^27^]. The scale has been previously used in the Indian context[^28,29^]. Anxiety was measured at time points T1 and T4. We found a strong internal consistency in our sample with a Cronbach’s alpha of 0.85 at T1.

#### Stress

The single item, *“Stress means a situation in which a person feels tense, restless, nervous or anxious or is unable to sleep at night because his/her mind is troubled all the time. Do you feel this kind of stress these days?* [^30^] was used to measure the level of stress experienced by the participants. The participants responded on a 5-point Likert ranging from *not at all* (1) to *very much* (5). We measured stress at T1 and T4.

#### Depressive symptoms

We used two items on depressive symptoms from the Patient Health Questionnaire-4 (PHQ-4) developed and validated by Kroenke et al. [^31^]to assess depressive symptoms. The scale included items such as *“Over the last two weeks, how often have you been bothered by the following: Feeling down, depressed or hopeless; Little interest or pleasure in doing things”* Responses were recorded in a 4 point Likert scale ranging from *not at all* (0) to *nearly every day* (4). A previous study has used this scale in the Indian population [^32^]. We collected data on depressive symptoms at T2 and T4.

### Predictors

#### Sociodemographic characteristics

Sociodemographic information of the study participants included age (in years), gender (male/female/ non-binary), education (high school or less/some college/above college), annual income (in Indian Rupees) (0-3,00,000 (low)/ 3,00,000-7,00,000 (medium)/ 7,00,000 and above (high)), and place of residence (rural/ urban). To reduce the survey length, we included a few sociodemographic variables in each of the four surveys.

#### History of mental health

We used the question, “*Have you suffered from depression or any mental health issues before*” to assess if the participant had any history of mental illness. The responses to this question were recorded as *yes* (1) or *no* (0).

#### Social support

We used the following two items to measure social support: *Is there someone you could count on to help you if you contracted the virus and got sick, for example, to take you to the doctor or help you with daily chores?*, and *If in these times due to unforeseen circumstances you need some extra help financially, could you count on anyone to help you, for example, by paying any bills, housing costs, medical expenses, or providing you with food or clothes?* Responses were converted to *yes* (1) or *no* (0).

#### Resilience

Resilience was assessed using the two-item brief Connor-Davidson Resilience Scale, developed by Vaishnavi, Connor & Davidson (2007)[^33^]. It includes items such as *“Are you someone who is: Able to adapt to change; Tend to bounce back after illness or hardship”* The participants responded on a 5 point Likert scale ranging from strongly disagree (1) to strongly agree (5). We collected data on resilience at T1. We found a moderate internal consistency of the scale in our sample (Cronbach’s alpha of 0.60).

### Other variables

#### Responsibility

We assessed, through self-reports at T2, whether there was any increase in responsibilities (social, financial, household, and personal) of the participants during the lockdown. The responses were recorded as yes (1) or no (0). The aggregate of the four responsibility scores reflected the total increased responsibility score.

### Statistical analyses

We first performed descriptive analyses to compute the distribution of outcomes at different time points across gender, relationship status, education, annual income, and place of residence. Next, we fitted separate linear two-level (observations nested within individuals) multilevel models for each outcome (anxiety, stress, and depressive symptoms) to assess the temporal changes in the outcomes. These models accounted for any autocorrelation of the responses from the same participants. Model 1 included the primary predictor time and the sociodemographic variables. Model 2 additionally adjusted for the interaction of gender with time. Model 3 further adjusted for the buffer factors social support and resilience, and a history of mental health issues. Additionally, we ran an ANOVA model to analyze the gender differences in scores of responsibilities during the lockdown. We set alpha at 0.05 in our study. All our models were run in STATA version 12 [^34^].

### Ethical considerations

The study was approved by the Institutional Ethics Committee, Indian Institute of Technology, Gandhinagar. All participants were informed about their voluntary participation through the introduction section in the Google and Microsoft forms. The participants were requested to read the details about the study, carefully read the instructions, and then respond to the survey. The participants were also informed that the collected data would be kept confidential and not be shared with anyone outside the research team. Email addresses of participants who gave consent for follow-up were collected as identifying information. Statistical analyses were performed on de-identified data.

## 3. Results

### 3.1. Preliminary results

We collected data on psychological outcomes from 159 Indian adults across a period of two months during the lockdown. Our sample comprised relatively young participants (mean age=27.44 years, SD=9.17 years). About 65% of the sample were men and the remaining were women. The annual income of about half of the participants was below 3,00,000 Indian Rupees (a cut-off representing an income allowing decent living in a one-bedroom apartment for a couple in most urban areas of India). About 55% of the participants were at least college-educated, while only about 11% of the participants reported having an educational qualification less than high school. The distribution of the psychological outcomes across these groups is presented in Table 1.

**Table 1:** Distribution (mean, standard deviation) of anxiety, stress, and depressive symptoms at T1, T2, and T4 across gender, annual income, education, and place of residence (N=159).

|  |  | Anxiety |  | Stress |  | Depressive symptoms |  |
| --- | --- | --- | --- | --- | --- | --- | --- |
|  |  | T1 | T4 | T1 | T4 | T2 | T4 |
| Gender |  |  |  |  |  |  |  |
| Men |  | 4.02<br>(4.41) | 4.17<br>(4.19) | 1.98<br>(0.90) | 2.29<br>(1.07) | 1.42<br>(1.27) | 1.67<br>(1.58) |
| Women |  | 4.61<br>(4.14) | 6.72<br>(4.83) | 2.36<br>(1.13) | 3.25<br>(1.22) | 1.59<br>(1.41) | 2.19<br>(1.58) |
| Annual income |  |  |  |  |  |  |  |
| Low |  | 4.59<br>(4.45) | 5.44<br>(4.94) | 2.10<br>(1.03) | 2.71<br>(1.20) | 1.57<br>(1.23) | 1.99<br>(1.62) |
| Medium |  | 4.37<br>(4.41) | 5.15<br>(4.59) | 2.18<br>(1.05) | 2.69<br>(1.36) | 1.64<br>(1.68) | 2.15<br>(1.81) |
| High |  | 3.30<br>(3.85) | 4.08<br>(3.57) | 2.08<br>(0.91) | 2.37<br>(1.05) | 1.31<br>91.02) | 1.24<br>(1.10) |
| **Education** |  |  |  |  |  |  |  |
| High school or less |  | 5.39<br>(5.86) | 4.17<br>(4.66) | 1.89<br>(0.76) | 2.78<br>(1.32) | 1.28<br>(0.96) | 1.5 (1.25) |
| Some college |  | 3.96<br>(4.12) | 6.02<br>(5.18) | 1.96<br>(0.90) | 2.77<br>(1.28) | 1.69<br>(1.35) | 2.35<br>(1.70) |
| Above college |  | 4.14<br>(4.05) | 4.63<br>(4.09) | 2.25<br>(1.09) | 2.60<br>(1.14) | 1.40<br>(1.36) | 1.62<br>(1.53) |
| **Place of residence** |  |  |  |  |  |  |  |
| Rural |  | 4.52<br>(3.65) | 6.38<br>(5.13) | 2.21<br>(0.98) | 2.75<br>(1.33) | 1.71<br>(0.95) | 1.96<br>(1.57) |
| Urban |  | 4.18<br>(4.42) | 4.81<br>(4.44) | 2.09<br>(1.00) | 2.6 (1.19) | 1.44<br>(1.37) | 1.83<br>(1.60) |

### 3.2 Trends of anxiety, stress, and depression during the lockdown

Our multilevel models adjusted for sociodemographic variables showed an increase in anxiety (**β**=0.81, CI: 0.03, 1.60) (Table 2) and stress scores (**β**=0.51, CI: 0.32, 0.70) (Table 3) during the two months (T1 to T4) of follow-up. We also found an increase in depressive symptoms (**β**=0.37, CI: 0.13, 0.60) in our sample between T2 and T4, independent of the covariates (Table 4). Anxiety (**β**=1.62, CI: 0.44, 2.81) and stress scores (**β**=0.65, CI: 0.37, 0.94) were found to be higher among women, versus men, independent of their sociodemographic factors. However, we could not find statistically significant associations of the other sociodemographic variables (age, annual income, educational qualification, and place of residence) with the psychological outcomes.

**Table 2:** Changes (regression coefficients, 95% CI) in anxiety between Week 1 and Week 8 of the lockdown (N=159).

|  | Model 1 | Model 2 | Model 3 |
| --- | --- | --- | --- |
| **Anxiety** |  |  |  |
| **Time** | 0.814 (0.025, 1.602) | 0.125 (-0.831, 1.082) | 0.168 (-0.860, 1.197) |
| **Gender (ref- Men)** |  |  |  |
| Women | 1.624 (0.443, 2.805) | 0.619 (-0.820, 2.059) | 0.306 (-1.170, 1.782) |
| Age | -0.042 (-0.113, 0.027) | -0.043 (-0.113, 0.027) | -0.055 (-0.125, 0.014) |
| **Annual income (ref-Low)** |  |  |  |
| Medium | 0.315 (-1.125, 1.756) | 0.286 (-1.155, 1.727) | 0.069 (-1.348, 1.486) |
| High | -0.734 (-2.178, 0.710) | -0.742 (-2.187, 0.702) | -0.694 (-2.145, 0.757) |
| **Education (ref- High school or less)** |  |  |  |
| Some college | 0.303 (-1.611, 2.217) | 0.320 (-1.594, 2.236) | 1.395 (-0.578, 3.370) |
| Above college | -0.399 (-2.393, 1.595) | -0.376 (-2.371, 1.618) | 0.777 (-1.243, 2.799) |
| **Place of residence (ref- Rural)** |  |  |  |
| Urban | -0.783 (-2.337, 0.769) | -0.790 (-2.343, 0.763) | -1.180 (-2.784, 0.423) |
| **Gender\*time** |  | 1.983 (0.357, 3.609) | 1.893 (0.155, 3.631) |
| **Social support** |  |  | -0.520 (-1.574, 0.533) |
| **Resilience** |  |  | -0.515 (-0.896, -0.135) |
| **History of mental health** |  |  | 2.236 (0.799, 3.673) |

**Model 1:** Adjusted for sociodemographic variables
**Model 2:** Includes interaction term (gender and time) adjusted for sociodemographic variables
**Model 3:** Additionally, adjusted for social support, resilience, and history of mental health

**Table 3:** Changes (regression coefficients, 95% CI) in stress between Week 1 and Week 8 of the lockdown (N=159).

| Stress | Model 1 | Model 2 | Model 3 |
| --- | --- | --- | --- |
| **Time** | 0.509 (0.315, 0.703) | 0.307 (0.073, 0.541) | 0.322 (0.075, 0.569) |
| **Gender (ref- Men)** | 0.654 (0.370, 0.938) | 0.362 (0.016, 0.709) | 0.270 (-0.092, 0.632) |
| **Age** | -0.009 (-0.025, 0.006) | -0.009 (-0.025, 0.006) | -0.011 (-0.027, 0.005) |
| **Annual income (ref- Low)** |  |  |  |
| Medium | 0.081 (-0.264, 0.428) | 0.081 (-0.264, 0.428) | 0.074 (-0.280, 0.429) |
| High | -0.083 (-0.428, 0.262) | -0.083 (-0.428, 0.262) | -0.056 (-0.418, 0.305) |
| **Education (ref- High school or less)** |  |  |  |
| Some college | 0.290 (-0.172, 0.752) | 0.290 (-0.172, 0.752) | 0.398 (-0.098, 0.896) |
| Above college | 0.282 (-0.196, 0.761) | 0.282 (-0.196, 0.761) | 0.375 (-0.131, 0.882) |
| **Place of residence (ref- rural)** |  |  |  |
| Urban | -0.062 (-0.435, 0.310) | -0.062 (-0.435, 0.310) | -0.213 (-0.613, 0.186) |
| **Gender\* time** |  | 0.583 (0.185, 0.980) | 0.577 (0.160, 0.994) |
| **Social support** |  |  | 0.025 (-0.238, 0.288) |
| **Resilience** |  |  | -0.063 (-0.158, 0.031) |
| **History of mental health** |  |  | 0.439 (0.085, 0.794) |

**Table 2:**
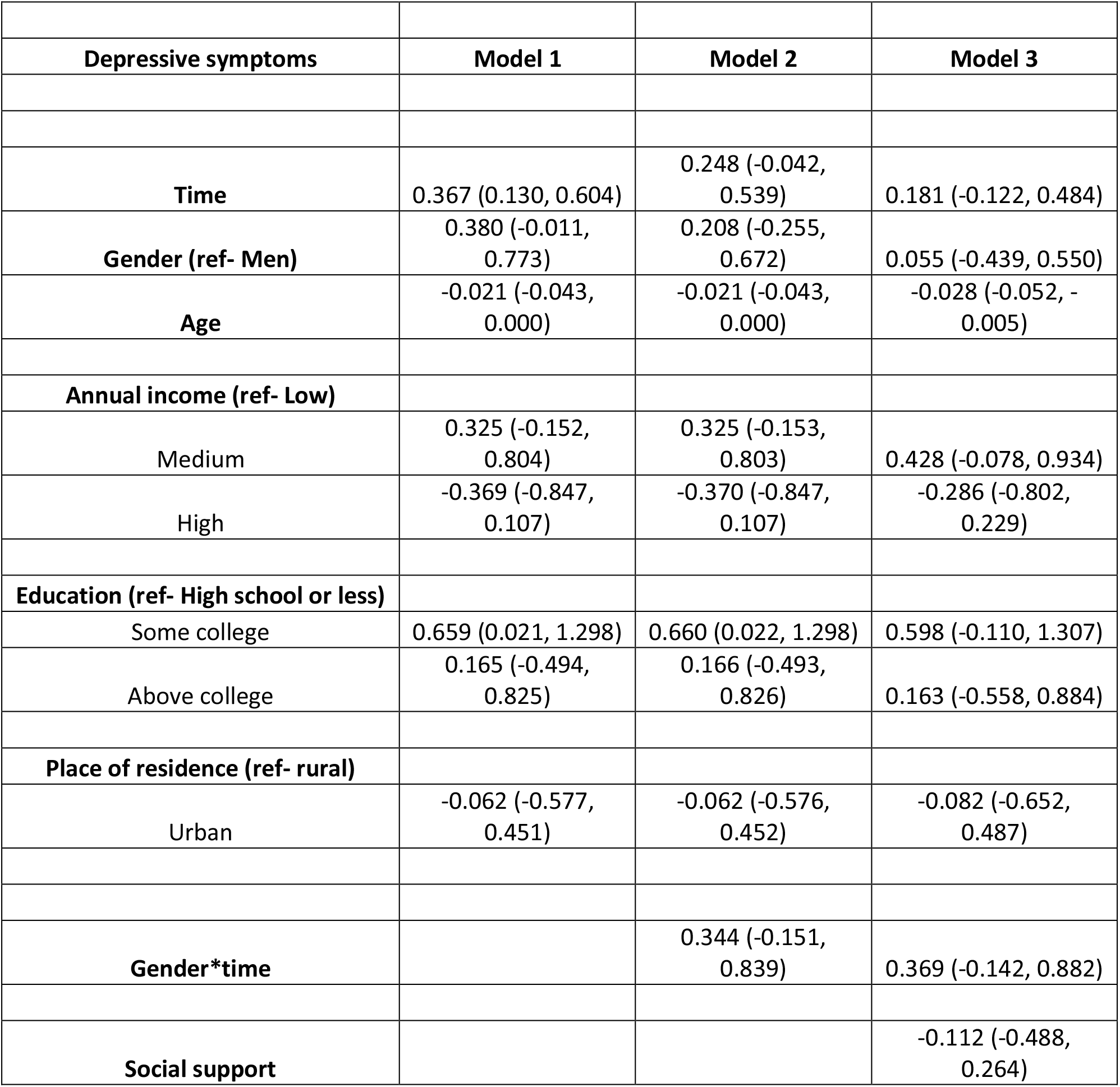
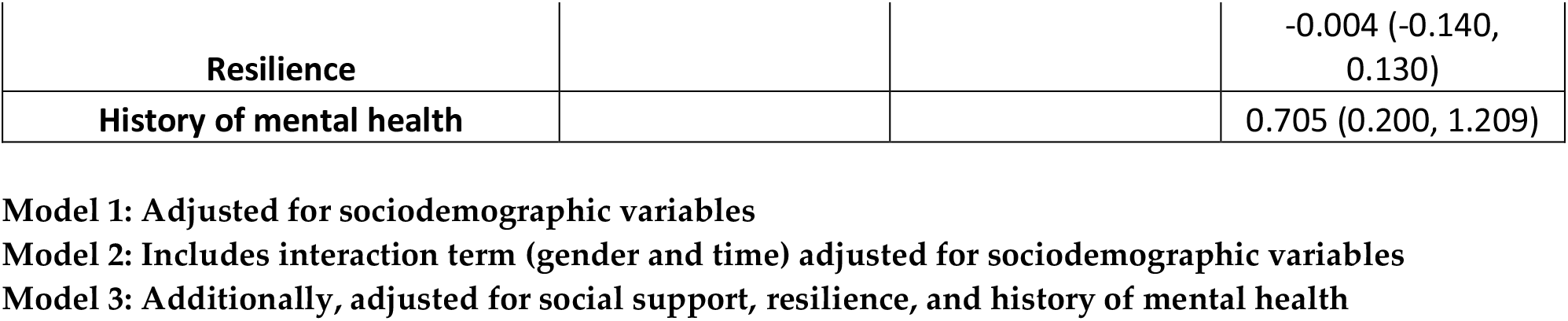
Changes (regression coefficients, 95% CI) in stress between Week 3 and Week 8 of the lockdown (N=159).

### 3.3 Differential increase of anxiety, stress, and depressive symptoms across gender

The interaction of gender with time was statistically significant for the anxiety (p-value for interaction = 0.017) and stress (p-value for interaction = 0.004) outcomes in our models adjusted for sociodemographic variables. Women showed a greater rate of increase in anxiety and stress scores between T1 and T4, as compared to men, after accounting for the sociodemographic covariates. Holding age, education, income and place of residence constant, men showed an average increase of 0.13 points in anxiety and 0.31 points in stress scores during the follow-up. The corresponding figures for women were 2.73 and 1.25 points, respectively. Further, this interaction was found to be significant even after adjusting for social support, resilience, and a history of mental health.

However, we did not find statistical evidence supporting the interaction of gender with time in our models for depressive symptoms.

### 3.4 Other findings

We found statistically significant and positive associations of history of mental health with anxiety (**β**=2.24, CI: 0.80, 3.67), stress (**β**=0.44, CI: 0.09, 0.79), and depressive symptoms (**β**=0.71, CI: 0.20, 1.21) in our sample, independent of all covariates.

Our fully adjusted models also found that a higher level of resilience was associated with lower anxiety, stress, and depressive symptoms. However, the associations of social support with the psychological outcomes were not statistically significant in our models.

Further, one-way ANOVA analysis highlighted a gendered difference in reports of increased responsibilities during the lockdown (p<0.001). Our results showed that women (M=1.35, SD=0.97) reported a greater increase in their responsibilities compared to men (M=0.90, SD=1.04) during the lockdown.

## 4. Discussion

Using repeated measures of psychological outcomes from 159 Indian adults during a period of two months of the Covid-19 lockdown, we found that there were statistically significant increases in stress, anxiety, and depressive symptoms over this period. Moreover, this increase in negative psychological outcomes was found to be more among women, compared to men. We also found that a higher level of an individual’s resilience was related to lower levels of anxiety, stress, and depressive symptoms.

Our findings suggest that anxiety, stress, and depressive symptoms increased during the lockdown among adults in India. While depressive symptoms increased in both genders, the effect size was modest. Nevertheless, the increase in the adverse psychological outcomes could be because of several reasons. First, the nationwide lockdown disrupted the citizens’ daily functioning in their professional, personal, and social lives, which potentially impacted their psychological well-being. Moreover, the periodic extensions of the lockdown over a considerably long period of time, accompanied by a steep increase in the number of Covid-19 cases in the country, and even worldwide likely worsened their anxiety, stress, and depressive symptoms over time. Each announcement of the extension of the lockdown might have increased the anxiety among the citizens by engendering a perception of unpredictability. Further, the initial shocks of the lockdown followed by the social isolation maintained for a prolonged time, and the emotional and financial losses incurred during the lockdown might have created a synergistic psychological impact. Second, the government of India announced a relaxation of restrictions on certain activities even while extending the lockdown. These relaxations included the resumption of trains, opening up of small shops, and inter-state mobility. However, these graded “unlocks” (which potentially allowed increasing physical mobility as well), were not accompanied by reports of a reduction in the number of new Covid-19 cases in India. On the contrary, the case numbers shot up just as the “unlocking” began. This could have reduced the confidence of the citizens, leading to a higher perceived risk of contracting the disease and thus further increasing their stress, anxiety, and depressive symptoms. However, these findings of our study are contrary to those found by Wang et al. [^35^] in China, where stress and anxiety were found to be stable across a 4-week period of lockdown among the Chinese people. The explanation provided by Wang et al. highlights that China recorded substantial improvements in curbing the spread of the virus through their rapid decisive measures and the greater number of recovered patients, which might have instilled greater confidence in their public health measures among the Chinese, thus avoiding a worsening of the psychological toll of a prolonged lockdown [^35^].

As per the findings of our study, compared to men, women had a greater increase in stress, anxiety, and depressive symptoms during the lockdown. Notably, the increases in anxiety and stress in our sample were primarily due to this evidence of a greater increase in anxiety and stress observed in women. There are two potential explanations for this finding. First, it was found that women in the Indian context have significantly more household responsibilities, during the lockdown especially because of the skewed gendered division of household labour in India [^36,37^]. For instance, due to the closure of schools and offices, all family members could be staying indoors leading to an increase in household burden for women who would be expected to shoulder most of the childcare, cooking, cleaning, other household management and coordination of other responsibilities in most households. This would leave them with a very limited time for themselves. Corroborating this, Viglione [^38^] reported lower publication rates among female academicians in North America compared to their male counterparts during this pandemic, across all disciplines. Our results from India, a society with stronger patriarchy, fit this narrative. We found that women reported a greater increase in their responsibilities during the lockdown compared to men. These added responsibilities combined with the lack of time for themselves could increase their stress, anxiety and depression levels much more than the increase among men. Second, social isolation and the restrictions on physical mobility might increase the exposure of women to hostility at home, especially among women who were already vulnerable to domestic violence. Previous studies have reported an increase in the risk of women across the world experiencing hostility during the lockdown period [^39–41^]. The high prevalence (∼30%) of domestic violence [^18^] in India is a reflection of the vulnerability of Indian women to domestic violence (ranging from emotional abuse from family members to physical/sexual abuse from intimate partners). Any risk of such hostility could worsen during the lockdown. Prolonged exposure to any risk of domestic hostility (emotional, physical or sexual) could lead to an increase in stress, anxiety, and depression among Indian women during the lockdown.

We found that the greater increase in stress and anxiety among women versus men persisted even after accounting for social support and resilience. This suggests that this gendered pattern was strong enough to persist despite any protective effect exerted by these buffering factors. Even though interaction of depressive symptoms with gender was not statistically significant, it suggested that the rate of increase in depressive symptoms was higher among women than men, thus fitting the pattern observed with anxiety and stress. Nevertheless, these findings are not in-line with the findings of a similar longitudinal study by Ozamiz et al. [^26^] in Northern Spain. They found higher levels of anxiety, stress, and depression among men compared to women. This contrast with our results could be because of the cultural differences between the two countries with regard to the gendered division of household labor [^42,43^].

We found that higher resilience likely dampened the increase of negative psychological outcomes among our participants. Resilience is known to buffer the impact of stress on mental health[^44^], especially during the Covid-19 pandemic[^45^]. Although not statistically significant, we also found evidence suggesting that social support reduced the intensity of increase in participants’ stress, anxiety, and depressive symptoms. This is in line with findings from previous research. It is likely that the initial announcement of the lockdown led to individuals moving to their natal/marital households or enhanced their interaction with their family members and loved ones, thereby increasing their social support[^4^]. Such support could act as an assurance of emotional, social, and financial challenges during the time of crisis. Such perceived assurance of help from others during the crisis could act as stress-ameliorating factors, preventing the increase of anxiety, stress, and depression. However, due to the small sample size, we did not have statistical power to support this.

We also found that persons with a history of mental health issues were likely to have an increase in anxiety, stress, and depression during the period of lockdown. Previous research has highlighted that situations of social avoidance could cause a relapse of trauma and depressive events[^46^]. The social isolation, the added responsibilities, and any lack of perceived social support (due to physical mobility restrictions) could trigger those with past depressive episodes. Studies have shown higher stress and anxiety during the lockdown in India [^47^].

We could not find statistical evidence to support the relationship of age, annual income, and education with anxiety, stress, and depressive symptoms in our sample. However, despite the lack of statistical significance, we found that a higher annual income, lower age, and living in urban versus rural areas, were related to lower levels of adverse psychological outcomes. A higher income could reflect employment that assured job security, flexibility, and a continued salary during the lockdown, which in turn may dampen the psychological impact of the prolonged lockdown. Also, it likely provides individuals with financial resources to better adapt to the crisis.

### Limitation and strengths

The study has several limitations which we acknowledge. First, the survey was conducted online limiting the sample to only those who had access to the Internet. However, the online method of recruitment helped us collect data from a diverse sample within a short time, given the restriction of physical mobility due to the lockdown. Second, we could not follow-up with the majority of our participants during the study. This was likely because we relied on only one way of communication, their email, for follow-up. In the chaos of the Covid-19 pandemic and the challenges it brought, the participants might have missed the emails related to the follow-ups. However, to our knowledge, this is the first web-based longitudinal study from India capturing key insights of psychological well-being *over time* during a lockdown that was periodically extended. Third, our sample size of 159 participants was modest; yet, it allowed us to analyze the changes in the psychological outcomes during the lockdown with several statistically significant results. Fourth, since the survey was in English, all participants who volunteered were comfortable in English and unsurprisingly 89% had some college education. Therefore, the results cannot be generalized to the whole of India. However, we found policy-relevant results showing an increase of anxiety, stress, and depressive symptoms in a relatively well-educated sample. We argue that the relatively underprivileged (socially as well as economically) are potentially even more vulnerable to such adverse psychological outcomes during the lockdown. Lastly, we used self-reported measures to assess anxiety, stress, and depressive symptoms in our sample. While clinical interviews would have yielded better results, we argue that the use of validated, reliable, and widely cited scales make our results credible.

Despite these limitations, the strength of our study lies in its longitudinal nature which sheds light on the trend of psychological outcomes during the lockdown in India. Moreover, we measured the outcomes at two interesting time points, one during the initiation of the lockdown and the other during a phase of relative relaxation, allowing us to assess if the psychological outcomes changed during differing dynamics of the lockdown. Despite a modest sample size, we also found statistical evidence to highlight the gender-based disparities in the effect of the lockdown, which was likely due to the gendered interpretation of circumstances created due to this pandemic plus a gendered emotional and behavioral response to the subsequent lockdown, all of which could in turn be socially determined.

### Implications

Our salient findings highlight a long-term impact of the lockdown on the mental wellbeing of Indian adults. These findings can help mental health policymakers to design disaster-response policies to address the psychological needs of the citizens during such crises, including a plan for follow-ups. Additionally, our findings suggest that these policies should be socially inclusive, with prioritized care for the vulnerable such as women and those with existing mental health issues. A longer-term perspective on preparedness would benefit from policies designed to enhance resilience among Indian citizens and prepare them to adapt to such crises.

An immediate response to our findings would be the involvement of philanthropic non-governmental organizations, social workers, and other community service providers to provide emotional support to communities during and after the Covid-19 pandemic, with a special focus on women and the underprivileged.

## 5. Conclusions

The Covid-19 crisis and the accompanying lockdown have no doubt affected everyone’s life in some way or the other. While the lockdown may help in effectively addressing this pandemic, the state and society at large need to be sensitive to the mental health impacts of a long-drawn out lockdown. Vulnerable populations such as women and the marginalized deserve immediate attention. However, our responsibility also lies in addressing the root causes driving the unequal distribution of psychological distress during such crises.

## Author Contributions

Conceptualization (initial), AG; Conceptualization (additional), AJS and MAS; methodology, AG and MAS; software, AJS; validation, AG and AJS; formal analysis, AJS; investigation, AG; resources, AG and MAS; data curation, AG; writing—original draft preparation, AG and AJS; writing—review and editing, AG, AJS, and MAS; supervision, MAS; project administration, AG and AJS. All authors have read and agreed to the published version of the manuscript.

## Data Availability

The data cannot be shared due to confidentiality promised to the participants

## Acknowledgments

This project was completed with the help and support of a lot of people, and we want to acknowledge their contribution. AG conveys special thanks to Vardhaman Gopal for his timely suggestion, which set the ball rolling and for being a pillar of support. AG is immensely grateful to her parents for being super helpful and patient during the entire process, particularly their unwavering support throughout the extended lockdown. AG thanks all her friends and family who not only helped in the circulation of the survey but were a continuous support. AG thanks Sitesh Kumar for his help with numerical data representation. AG, AJS, and MAS thank Nilesh Thube for his help in the results tables. We cannot thank the participants enough for their continuous contribution towards diligently filling up the forms longitudinally. AG was supported by fellowship from University Grants Commission (UGC)-NET-JRF, India. AJS was supported by fellowship from the Department of Science and Technology, India and IIT Gandhinagar.

